# Yin-Yang 1, a player regulated systematic inflammatory involved in cognitive impairment of depression

**DOI:** 10.1101/2022.06.19.22276593

**Authors:** Jing Lu, Kangyu Jin, Jianping Jiao, Ripeng Liu, Tingting Mou, Bing Chen, Zhihan Zhang, Chaonan Jiang, Haoyang Zhao, Zheng Wang, Rui Zhou, Manli Huang

**Author notes:** The corresponding authors contribute equally to this work, **Corresponding author:** Rui Zhou, PhD., Affiliation: Department of Nuclear Medicine and Medical PET Center, The Second Affiliated Hospital of Zhejiang University School of Medicine, Hangzhou, 310009, China, Manli Huang, M.D., Affiliation: Department of Psychiatry, The First Affiliated Hospital, Zhejiang University School of Medicine; the Key Laboratory of Mental Disorder’s Management of Zhejiang Province, Hangzhou 310003, China. The authors contribute equally to this work (Jing Lu, Kangyu Jin).

## Abstract

A growing number of clinical and preclinical studies suggest that alterations in peripheral and brain immunal system and followed inflammation are associated with the pathophysiology of depression, also leading to the changes in local glucose metabolism in the brain. Here, we identified Yin-yang 1 (YY1), a transcription factor that has been reported to be closely associated with central and peripheral inflammation. The levels of YY1 and IL-1β were significantly increased in blood samples from depressed individuals, and significantly decreased after treatment with Vortioxetine. Meanwhile, it was found that the level of YY1 in plasma was negatively correlated with visual learning reasoning and problem solving in MDD patients, and positively correlated with the level of IL-1β in plasma. CUMS animals showed depressive-like behavior. Compared with the control group, MicroPET analysis showed that the decrease of glucose metabolism in the hippocampus, entorhinal cortex, amygdala, striatum and mPFC was reversed after treatment. After treatment, these changes were reversed. In conclusion, Our study suggested that YY1-NF-κB - IL-1β inflammatory pathway may play an essential part on both mood changes and cognitive impairment in depression, and may be associated with changes in glucose metabolism in the emotion regulation and cognition related brain regions. These findings provide new evidence for the inflammatory mechanisms of depression.

## 1. Introduction

Major depressive disorder (MDD), is a severe affective disorder characterized by a group of symptoms including persistent low mood, worthlessness or guilt, anhedonia, inattention, changes in appetite, mental slow movement or agitation, sleep disturbance, or suicidal thoughts, imposing a heavy social burden(Karrouri et al., 2021). Depression is often accompanied by the clinical symptom of cognitive impairment, which resulting in a decline in quality of life and psychosocial function(Frampton, 2016). The DSM-5 has defined cognitive impairment as one of nine diagnostic criteria for depression, including impaired thinking, memory, attention and decision making. Cognitive dysfunction in MDD mainly displays in learning, attention, memory, processing speed and executive function (Varghese et al., 2022, Bortolato et al., 2016). Because of its potential contribution to the full recovery of function in MDD patients, the remission of cognitive dysfunction is increasingly being promoted as an important goal in the treatment of major depression(Lam et al., 2014). Additionally, increasing studies have shown that cognitive disorders may have complex interactions with emotional symptoms. For example, the aggravation of depressive symptoms may negatively affect cognitive function(McDermott and Ebmeier, 2009)and the changes in cognitive function may be related to the response of patients to antidepresant treatment(Culang et al., 2009). Therefore, the study of the pathophysiological mechanism of cognitive impairment in depression is conducive to further understanding the pathogenesis of depression, and may find new therapeutic targets for clinical diagnosis and treatment.

Transcription factor Yin Yang 1 (YY1), firstly identified in 1991, is widespread expression in mammals. Previous studies showed that YY1 plays an important role in early neurodevelopment(Shiu et al., 2016),(Dong and Kwan, 2020),(Zambrano et al., 1997). Some recent studies revealed that YY1 also plays a role in inflammation regulation (Darcy et al., 2020). Zhang et al. found that YY1 was gradually upregulated in lipopolysaccharide (LPS)-stimulated BV2 microglia, which was dependent on the transactivation function of nuclear factor kappa B (NF-κB). Furthermore, LPS-induced activation of NF-κB signaling pathway and increase of interleukin 6 (IL-6) expression were significantly inhibited in microglia of YY1 knockout mice. Meanwhile, YY1 enhanced the binding of NF-κB to the IL-6 promoter, reducing the H3K27ac modification of the IL-6 promoter, and finally increasing IL-6 transcription(Zhang et al., 2018). Our previous study found that YY1 expression was increased in the peripheral blood mononuclear cell (PBMC) from MDD patients in single-cell RNA sequencing analysis compared with healthy volunteers(Lu et al., 2021). These studies suggested that YY1 is closely associated with both central and peripheral inflammation. Inflammation was proved to be associated with depression and cognitive impairment. Reichenbergde et al. demonstrated that cytokines could mediate affective and cognitive disturbances in humans, intravenous administration of S. equinum endotoxin significantly increased circulated TNF-α, soluble TNF receptors, IL-6 and IL-1 receptor antagonists in healthy volunteers, which elevated the level of depressed mood and impaired cognitive functions such as verbal memory and non-verbal memory(Reichenberg et al., 2001). Many other studies have also reported that inflammation may be attributed to depression and its cognitive impairment(Zhao et al., 2022, Jin et al., 2020) Thus, to investigate the role of YY1 in depression and cognitive impairment is of great importance.

Examination of the brain using positron emission tomography (PET) may lead to a better understanding of the underlying pathophysiology of MDD, which makes it possible to demonstrate changes in the regional brain metabolic rate (rCMRGlu) of glucose in vivo(Hendler et al., 1997). Recent studies have shown that brain hypometabolic pattern is closely related to the type and severity of cognitive deficits(Laforce et al., 2018). The roles of peripheral inflammation on glucose metabolism in the parahippocampal and rhinal directly mediate the effects of inflammation on spatial memory impairment(Harrison et al., 2014). However, little is known about the changes of brain glucose metabolism before and after treatment in depression animal model and its relationship with inflammation and behaviour.

Vortioxetine is a novel multimodal antidepressant with 5-HT receptor modulator and serotonin transporter (SERT) inhibitor properties(Pehrson et al., 2018). A number of preclinical and clinical data have demonstrated the antidepressant effects of Vortioxetine(Alvarez et al., 2012, Mork et al., 2012), and many preclinical studies have suggested that vortioxetine can improve certain aspects of MDD related cognitive impairment, such as processing speed, executive function, and memory(du Jardin et al., 2014, Li et al., 2015). Baune et al. also reported that vortioxetine had beneficial effects on mood and cognitive function of patients with major depression(Baune et al., 2018). It is noteworthy that vortioxetine may have anti-inflammatory effects due to its structural similarity to non-steroidal anti-inflammatory drugs(Bayram et al., 2018). Furthermore, in one study of human monocytes/macrophages, vortioxetine showed antioxidant activity and anti-inflammatory effects, polarizing macrophages into other phenotypes(Talmon et al., 2018). In addition, the antidepressant effects of vortioxetine were enhanced by the anti-inflammatory celecoxib(Fourrier et al., 2018). Vortioxetine may, contrary to what was previously believed, play an antidepressant and improve cognitive function in part through its anti-inflammatory effects, Therefore, it is necessary to study the immunomodulatory mechanism of Vortioxetine in depressed individuals.

Therefore, this study aims to elucidate the changes of YY1-related inflammatory pathways before and after treatment of depression and its relationship with cognitive function. First, we measured the level changes of proinflammatory factors, YY1 and cognitive function in peripheral blood of MDD patients before and after treatment, and explored their correlations. Second, three groups were constructed, including control, CUMS and VOR groups, and the changes of behavior, glucose metabolism and mRNA level of YY1 and proinflammatory factors were detected. In conclusion, our study reveals the role of YY1-related inflammatory pathway in MDD and cognitive impairment, and provides theoretical support for seeking new targets for clinical diagnosis and treatment.

## 2. Material and Methods

### 2.1 Subjects

Of the original 90 adults (45 MDD patients, 45 healthy controls (CTR)), 3 patients fell out at follow-up after eight weeks of treatment, and 2 CTR participated in the scale evaluation but refused to have their blood drawn. Finally, a total of 80 Han right-handed adults participated in this study, including 40 first episode, medicine-naïve MDD patients (aged 18-45 years old) and 40 age, sex and education CTR. MDD patients were recruited from the psychiatric department of First Affiliated Hospital, College of Medicine, Zhejiang University. MDD patients were treated by vortioxetine (10-20mg) for 8 weeks. Subjects characteristic and symptoms assessment were concluded in Table 1-2. The inclusion criteria of MDD were as follows: (1) age 18–45 years; (2) meet the Diagnostic and Statistical Manual of Mental Disorders, IV Edition (DSM-IV) criteria for first-episode current unipolar MDD; (3) score of 17 items of Hamilton Depression Scale (HAMD-17) ≥17; (4) with at least 9 years of formal education; (5) no alcohol or substance abuse. Healthy volunteers matching age, sex and education level are recruited from the local community through advertising. Exclusion criteria for all participants were as follows: (1) MDD patients who had received any form of treatment prior to the study; (2) major medical diseases; (3) organic brain disease; (4) other psychiatric disorders (except major depressive disorder); (5) dementia or other cognitive impairment; (6) pregnant or planning to become pregnant recently; (9) an active medical condition that may be associated with depressive episodes (e.g., untreated hypothyroidism, psoriasis and uncontrolled cardiovascular events in the past 6 months).

**Table 1.**
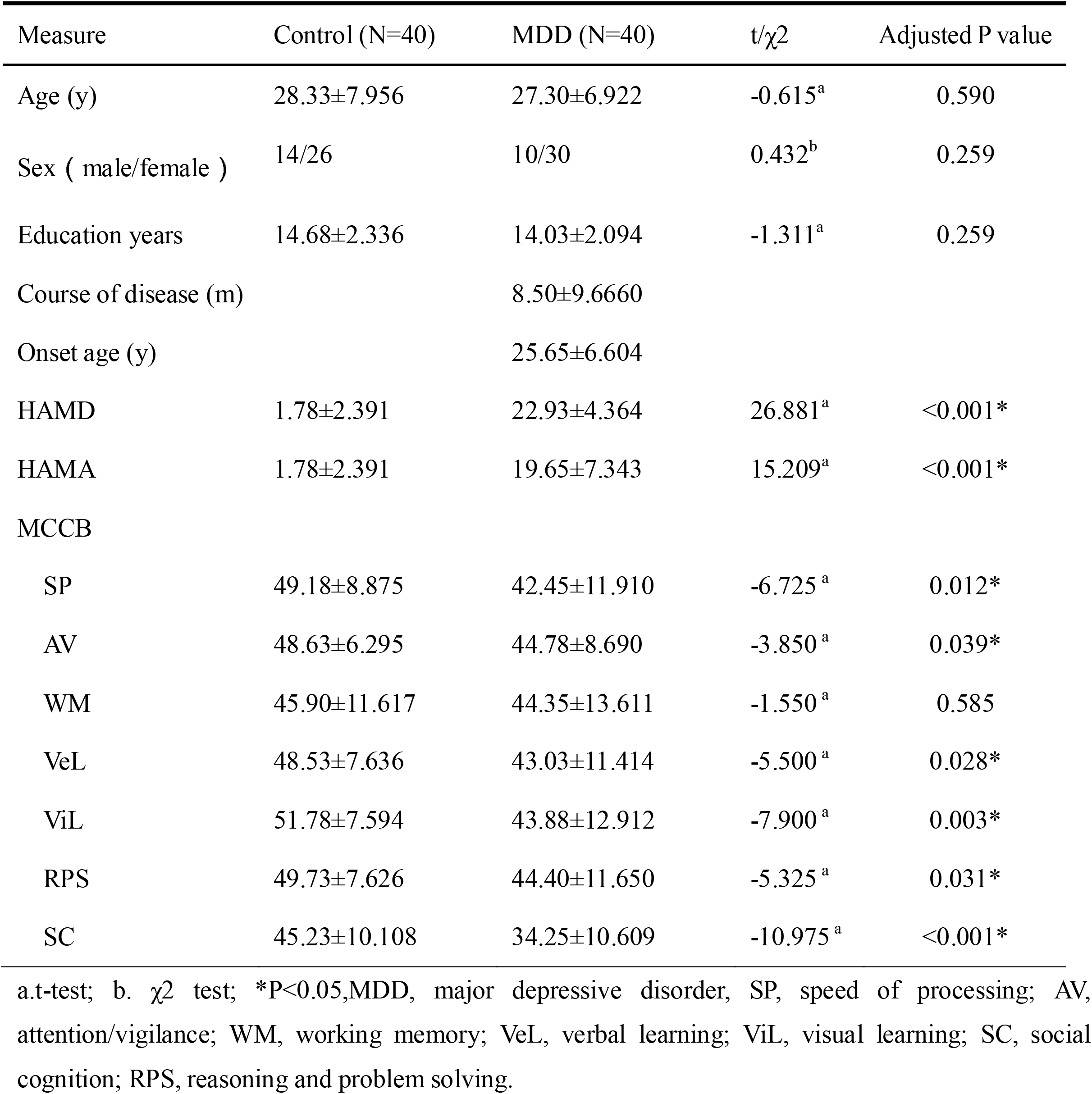
Subjects Characteristic

**Table 2.** Symptoms assessment of MDD and after vortioxetine treatment

| Measure | MDD (N=40) | VOR (N=40) | t/$\chi^2$ | Adjusted P value |
| --- | --- | --- | --- | --- |
| HAMD | 22.93±4.364 | 10.45±6.320 | 12.227<sup>a</sup> | <0.001* |
| HAMA | 19.65±7.343 | 9.63±6.229 | 7.530<sup>a</sup> | <0.001* |
| MCCB |  |  |  |  |
| SP | 42.45±11.910 | 48.97±11.384 | -3.956<sup>a</sup> | <0.001* |
| AV | 44.78±8.690 | 49.36±9.842 | -4.301<sup>a</sup> | <0.001* |
| WM | 44.05±13.655 | 45.67±10.322 | -0.717<sup>a</sup> | 0.478 |
| VeL | 43.44±11.260 | 50.36±11.056 | -4.292<sup>a</sup> | <0.001* |
| ViL | 43.90±13.080 | 51.69±9.459 | -5.790<sup>a</sup> | <0.001* |
| RPS | 44.31±11.788 | 49.21±10.136 | -3.968<sup>a</sup> | <0.001* |
| SC | 34.23±10.747 | 35.87±13.215 | -0.921<sup>a</sup> | 0.408 |

a. paired t-test; b. $\chi^2$ test; \*P<0.05, MDD, major depressive disorder, VOR, vortioxetine treatment group, SP, speed of processing; AV, attention/vigilance; WM, working memory; VeL, verbal learning; ViL, visual learning; SC, social cognition; RPS, reasoning and problem solving.

All subjects knew the content of the trials and signed informed consent. The study was approved by ethics review boards of the First Affiliated Hospital of Zhejiang University School of Medicine.

### 2.2 Symptoms assessment

HAMD-17 consists of five components of anxiety/body mass, weight, cognitive impairment, retardation, and sleep disorders, with a total of 17 items used to assess depressive symptoms. Anxiety is measured using the Hamilton Anxiety Scale (HAMA), which includes two components: mental and physical anxiety. We tested the visual learning (ViL), reasoning, and problem solving (RPS) ability of subjects using the standardized measurement tool MATRIX Consensus Cognitive Battery (MCCB). MCCB has been shown to have cognitive functions such satisfactory confidence in assessing patients’ schizophrenia and depression cognition(Jin et al., 2020) (Buchanan et al., 2011).

### 2.3 Plasma cytokine levels detection

Approximately 3-5 ml of blood was aseptically collected from each patient using an approved standard procedure. Subsequently, the blood samples were centrifuged at 3,000 rpm for 15 minutes followed by separated and stored at -80 °C before analysis. The plasma IL-1β, IL-6 and YY1 levels were assessed using ELISA kit (Human IL-1β, HSLB00D, R&D; Human IL-6, HS60DC, R&D; Human YY1, SEF572Hu, Cloud-Clone Corp.).

### 2.4 Experimental animals

The 24 healthy Sprague Dawley (SD) male rats (8 weeks old) were randomly assigned into control group (n = 8) and chronic unpredictable mild stimulation (CUMS) (n = 16). CUMS protocol was consistent with our previous study and consisted primarily of a 8-week daily exposure to alternating stressors along with occasional nighttime stressors(Lu et al., 2017). All CUMS rats were randomly divided into two groups after the modeling successfully that was confirmed by forced swimming test and sugar water preference test(Wang et al., 2021): CUMS, vortioxetine treatment (VOR) group. Eight CUMS rats were treated with Vortioxetine (10 mg/kg, Lu AA21004 hydrobromide, MedChemExpress) solubed in 1% DMSO for 4 weeks. Behavior test were conducted after 4 weeks treatment.

### 2.5 Behavioral assessments

Sucrose preference test (SPT), elevated plus test (EPT), open field test (OFT) and forced swimming test (FST) were used to assess depressive and anxiety-like behaviors. Learning and memory behaviors were assessed by Y-maze and Morris water-maze according to previous protocols (Ji et al., 2019, Guan et al., 2021).

### 2.6 Real-time quantitative PCR (RT-qPCR) for mRNA expression

The RNA expression levels of YY1, IL-1β, IL-6, NF-kB were detected by using RT-qPCR according to previous protocols (Lu et al., 2015). Briefly, commercial RNA isolation kits (EZBioscience,USA) were used to collect total RNA from the brain tissue. 4×EZscript Reverse Transcription Mix II kit (EZBioscience,USA) was used to synthesize cDNA. RT-qPCR was performed using 2× Universal SYBR Green Fast qPCR Mix (RK21203, ABclonal, China) in a CFX384 Touch Real-Time PCR Detection System(Bio-Rad, USA).All procedures were conducted following the manufacturer’s instructions. The expression levels of individual genes have been normalized to β-actin expression, and then calculated by using the 2^−ΔΔCt^ method. All primer pairs were designed with the program Primer Premier 5.0 (see Table 3).

**Table 3.**
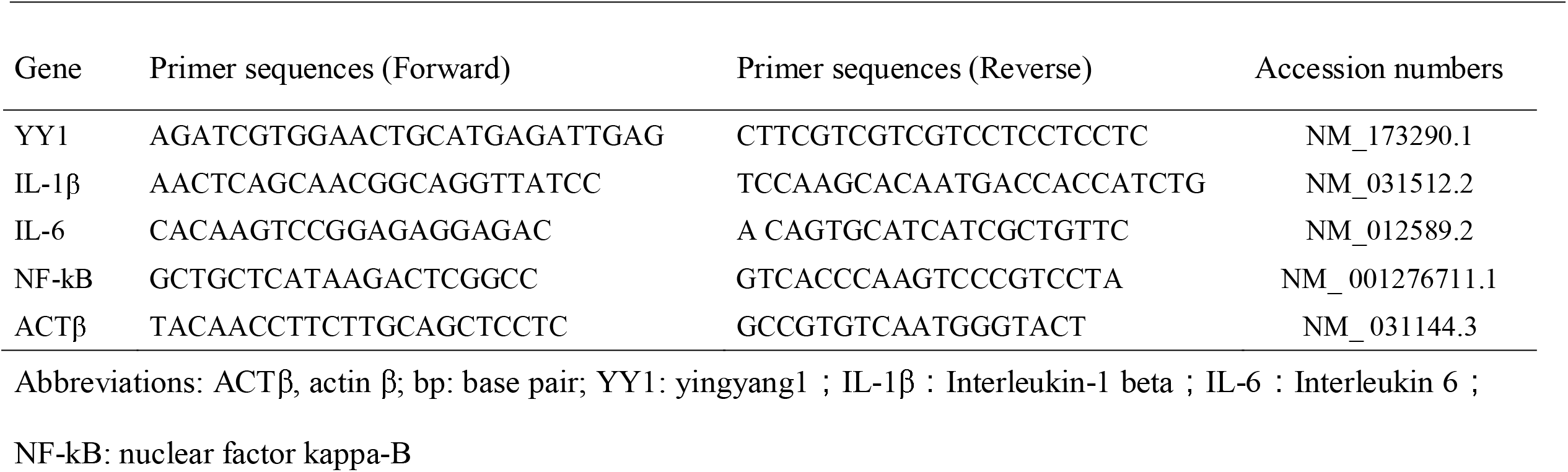
Sequence of the primers, gene bank accession numbers for the target genes and reference gene

### 2.7. Western blot assay for protein expression

Cytoplasmic and nuclear proteins were extracted from the hippocampus using a nuclear and cytoplasmic protein extraction kit (Beyotime.P0028), followed by BCA protein concentration determination Kit (Beyotime, P0026). The extracted protein supernatant was mixed with 5 × SDS-PAGE loading buffer, and the protein sample was prepared by boiling water bath for 15 min. 20.0 μg of protein sample was added to each line. After electrophoresis for 1.0 h and transference to polyvinylidene fluoride membranes, the membrane was blocked in blocking buffer (with 5% bovine serum albumin) and incubated with primary antibodies overnight on a 4°C shaker. The primary antibodies we used were as follows: anti-P65(AF5006, 1:1000, Affinity Biosciences), anti-TBP(66166-1-lg,1:2000, Proteintech), anti-Beta Actin (66166-1-lg,1:1000, Proteintech). After washing, the membrane was incubated with the corresponding secondary antibody and colorified with Thermo Fisher Scientific. The blotting was detected by ECL Chemiluminescence Kit (Servicebio).

### 2.8 ^18^F-FDG microPET/CT scanning and image analysis

The energy source of brain is almost from glucose. Glucose metabolism in the brain is associated with the emotional and cognitive function. ^18^F-FDG, as the analogue of glucose, is widely used to noninvasively reflect glucose metabolism. Therefore, we used ^18^F-FDG to investigate glucose metabolic changes of brain regions associated with emotion and cognition in CUMS group and vortioxetine treated CUMS (VOR group). 18-fluorine fluorodeoxyglucose (^18^F-FDG) microPET/CT scanning was conducted by using a high-resolution small animal PET/CT (SuperArgus, Spain). Prior to ^18^F-FDG scanning, rats were fasted for 8 h, followed by anesthesia with 5% isoflurane and maintenance of anesthesia with 3% isoflurane. Static PET images were acquired for 10 min after 20 min of 18.5 MBq ^18^F-FDG (500 µCi) intravenous injection. Followed by PET scanning, a CT scan was performed to collect anatomic information. PET images (128 × 128 matrix, pixel size: 0.7764 × 0.7764 × 0.7963 mm) were reconstructed by using 3-dimensional ordered-subset expectation maximization (OSEM3D) algorithms.

PET and CT images were co-registered by using PMOD software (version 3.902, PMOD Technologies Ltd.) (Du et al., 2019). Fusional PET/CT images were co-registered with rat brain template (W. Schiffer) enclosed in PMOD software. To perform semiquantitative analysis, the volume of interest (VOI) of brain regions including hippocampus (HIP), amygdala (AMY), striatum (STR), entorhinal cortex (EC)and median prefrontal cortex (mPFC), were automatically drawn by PMOD software. Pons was selected as the reference region. Standard uptake value ratio (SUVR) was calculated by the following formula: SUVR = SUV (interest brain regions) / SUV (pons).

### 2.9 Statistical analysis

The data are presented as means * standard deviations for variables that are normally distributed, or as medians * quartiles for variables that are non-normally distributed. Missing data is estimated from the median in the given group. The Shapiro-Wilk test is used to determine the normal distribution. Mann-whitney U test was used to determine non-normal distribution variables between the two groups, T-test and one-way ANOVA were used to determine significant differences in normal distribution variables, and Dunnett-t test was used for post hoc analysis. The Chi-square test is used to classify variables. The Benjamini & Hochberg method is used to adjust P values in multiple tests. Pearson correlation index was used to test the correlation between the two groups.

## 3. Result

### 3.1 Clinical and demographic characteristics of subjects

The clinical and demographic characteristics of sujects are presented in Table 1. There was no significant difference either in gender, age or education years between MDD and CTR group. MDD and CTR subjests showed significant difference in HAMD and HAMA scores (all p< 0.001). And MDD patients were also observed siginificant lower MCCB scores in SP, AV, VeL,ViL, RPS and RPS than the CTR group. MDD patients showed significant changes in the symptoms assessment include HAMD and HAMA scores and SP, AV, VeL, ViL, RPS in MCCB assessment (all p< 0.001) after treatment of vortioxetine (see Table 2).

### 3.2 Cytokine characteristics of subjects

To investigate level of plasma YY1 and inflammatory factors, peripheral plasma of MDD patients and healthy control was collected to perfrom ELISA assay. The result indicated that plasma YY1 level increased in MDD patients (0.4982 ± 0.2836 ng/ml) compared with CTR(0.0274 ± 0.0215 ng/ml, p<0.001) and reduced after treated with vortioxetine (0.2279 ± 0.1969 ng/ml, p<0.001) (Figure 1A). Plasma level of IL-1β was increased in MDD (565.4724 ± 405.3974 pg/ml) than CTR (371.5978 ± 153.6342 pg/ml, p<0.001) and reduced after treatment ((187.9717±104.7408 pg/ml, p<0.001, Figure 1B). Plasma level of IL-6 was increased in MDD (1287.4829±789.9515 pg/ml) than CTR (608.2849 ± 506.9919 pg/ml, p<0.0001) while showed no significant change after treatment (912.2897 ± 707.8910 pg/ml Figure 1C, p=0.547).

**Figure 1.**
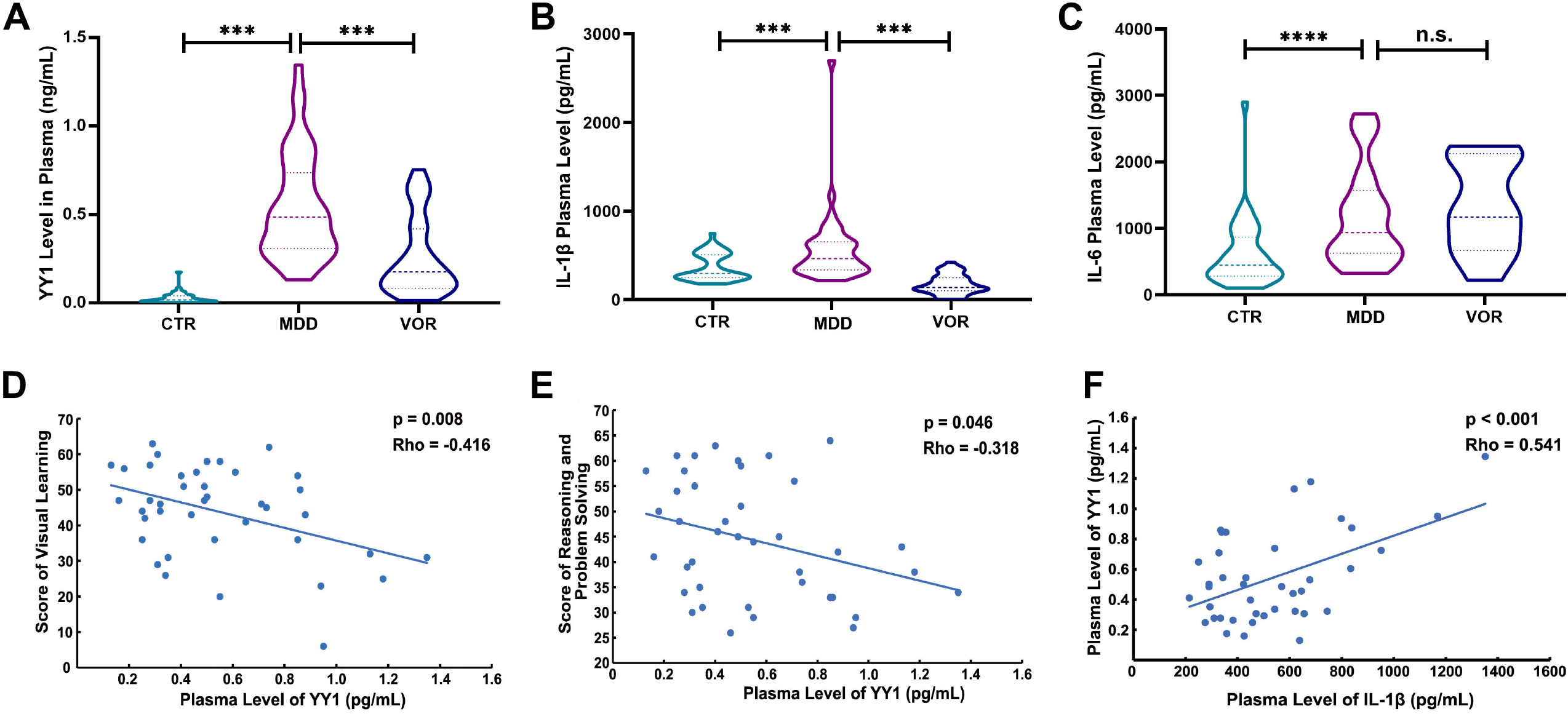
Changes of plasma level of Yin Yang 1 (YY1), IL-1β and IL-6 levels in patients with major depressive disorder (MDD, n = 40) and healthy controls (CTR, n = 40). (A) Comparison of YY1 in MDD and CTR. (B) Comparison of IL-1β in MDD and CTR; (C) Comparison of IL-6 in MDD and CTR; (D) Correlations between plasma YY1 levels and score of visual learning; (E) Correlations between plasma YY1 levels and score of reasoning and problem solving in MDD; (F) Correlations between plasma YY1 levels and plasma level of IL-1β in the patients with MDD. (*p < 0.05, **p < 0.01, ***p < 0.001, **** p<0.0001)

In order to determine whether the plasma level of YY1 was associated with cognition and cytokines, Pearson correlation analysis was conducted in MDD patients. The result showed that plasma YY1 was negatively correlated with the score of visual learning (p = 0.008, rho = -0.416, Fig. 1D) and the score of reasoning problem solving (p = 0.046, rho = -0.318, Fig. 1E), Plasma YY1 level was positively correlated with plasma IL-1β level (p < 0.0001, rho = 0.541, Fig. 1F). while these correlations were absent in control (all p > 0.05).

### 3.3 Behaviour and Cytokine characteristics of rats

The CUMS rats showed significant anxiety-like behavior in open field test (Fig. 2B-D), which showed significant decrease total distance in CUMS group in comparison to CTR and VOR group (p=0.0005, p=0.1777, respectively) and in center zone entries(p<0.0001 and p=0.0247, respectively). Depression-like behavior also performed, Sucrose preference test (CUMS vs. CTR p=0.0004, CUMS vs. VOR p=0.0034, Figure 2M) and forced swim test (CUMS vs. CTR p=0.0001, CUMS vs. VOR p=0.0020, Figure 2L) and and learning and memory behavior test (Elevated plus maze, CUMS vs. CTR p=0.0095, CUMS vs. VOR p=0.0079, Figure 2H-I; Y-maze, CUMS vs. CTR p=0.0398, CUMS vs. VOR p=0.0033, Figure 2J-K; Morris water maze, CUMS vs. CTR p=0.0001, CUMS vs. VOR p=0.0008, Figure 2E-G), and VOR treatment could partly reverse these performance.

**Figure 2.**
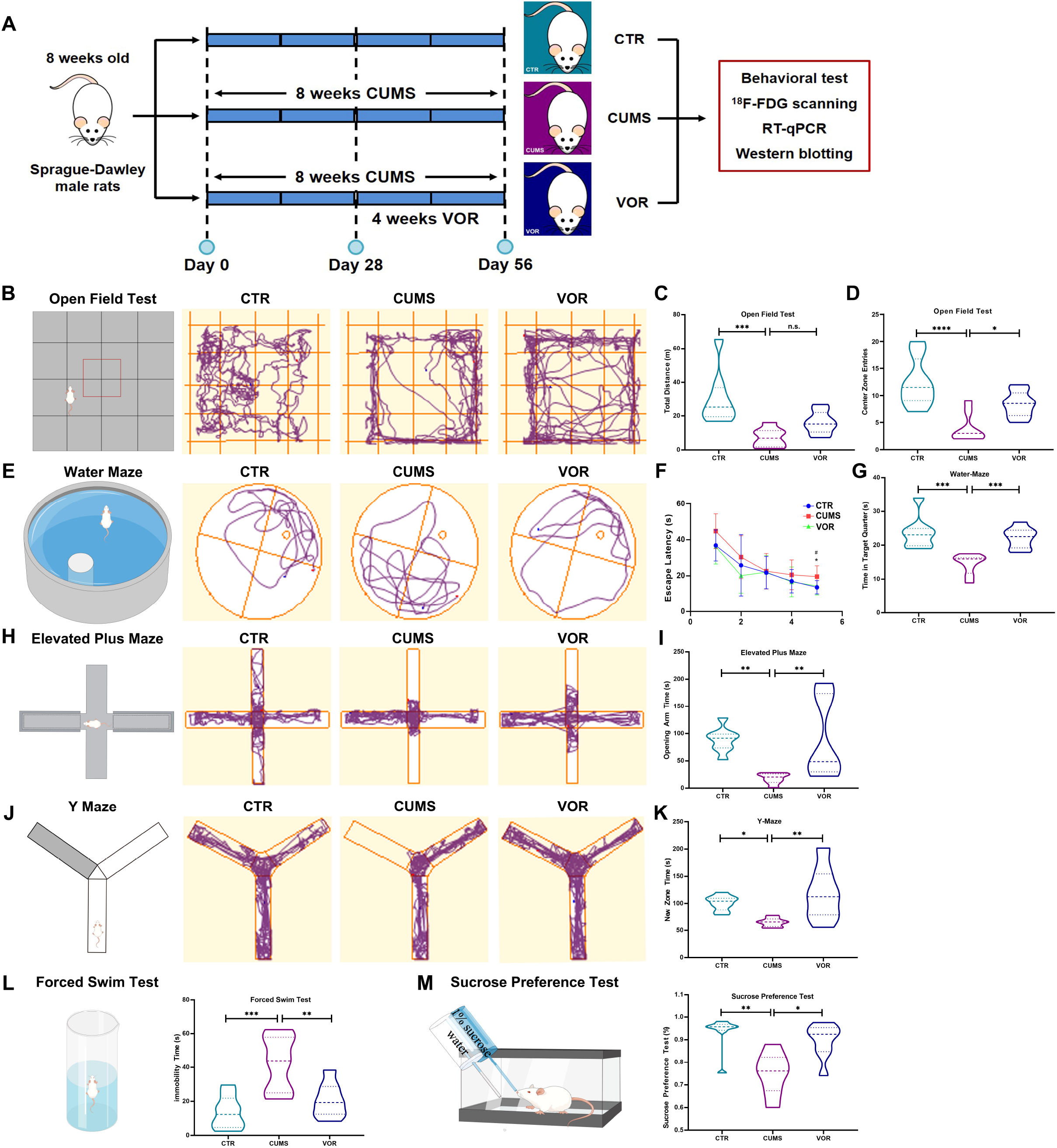
Vortioxetine treatment ameliorates depression-like and cognitive dysfunction behaviors in CUMS rat. (A) Diagram of experimental schedule; (B-D) Open field test. Trajectories of open field test (B); Total distance in open field test (C); Center zone entries in open field test (D). (E-G) Morris water maze test. Representative swimming paths during the spatial probe test (E); The latency to the platform of 5-day acquisition training in Morris water maze test (F, * means CTR vs. CUMS; # means VOR vs. CUMS); (G) The time in target quarter. (H and I) Elevated plus maze. Representative swimming paths during elevated plus maze (H). The time spent in open arm (I); (J and K) Y maze test. The representative swimming paths during Y maze test (J); the time spent in the new arm during the test (K); (L) Forced swim test; (M) Sucrose preference test. (n = 7 per group, ^#^ p<0.05, *p < 0.05, **p < 0.01, ***p < 0.001, n.s.: non-significant difference)

### 3.5 Glucose metabolism in brain regions of CUMS rats

Altered glucose metabolism in mPFC, striatum, amygdala, hippocampus and entorhinal cortex were observed (Fig. 3A). Tilted frontal-side view showed the location of altered brain regions (Fig. 3B). The results indicated that glucose metabolism of left mPFC (p=0.0102, Fig. 3C), striatum (p=0.0001, Fig. 3D), amygdala (p=0.019, Fig. 3E), hippocampus (p=0.0045, Fig. 3F) and entorhinal cortex (p=0.0006, Fig. 3G) was significantly decreased in CUMS compared with that of CTR group, and decreased in amygdala (p=0.0003, Fig. 3E), hippocampus (p=0.001, Fig. 3F) and entorhinal cortex (p=0.0262, Fig. 3G) after VOR treatment, while no significant changes in left mPFC and left striatum after VOR treatment (p=0.2569, p=0.088, respectively). CUMS rats showed significantly decrease glucose metabolism than controls and elevated after VOR treatment in the right mPFC (p<0.0001, p=0.0122, respectively, Fig. 3C), striatum (p=0.0245, p=0.0510, respectively, Fig. 3D), amygdala (p=0.012, p=0.01, respectively, Fig. 3E), hippocampus (p=0.0029, p=0.0030, respectively, Fig. 3F) and entorhinal cortex (p=0.003, p=0.006, respectively, Fig. 3G)

**Figure 3.**
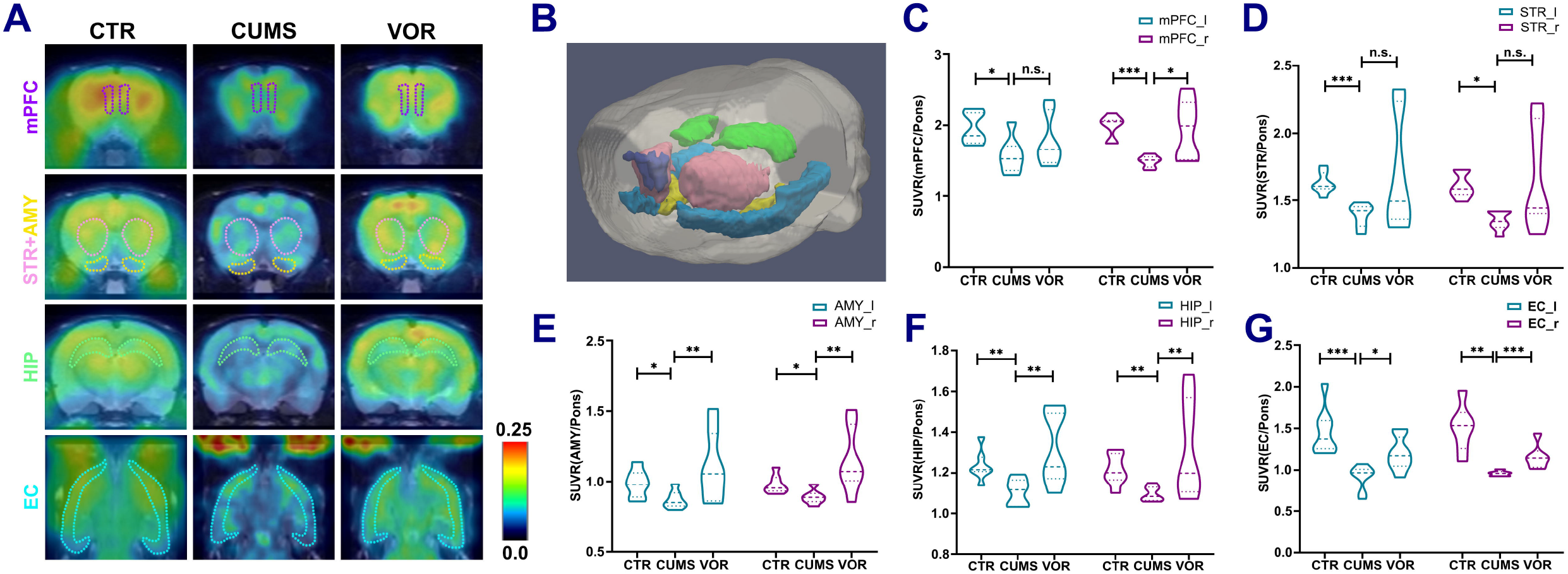
The glucose metabolic changes of hippocampus (HIP), amygdala (AMY), striatum (STR), median prefrontal cortex (mPFC) and entorhinal cortex (EC) in control group (CTR), depression model (CUMS group) and antidepressant (vortioxetine) treated group (VOR group). (A) Representative ^18^F-FDG PET images of HIP, AMY, STR, mPFC and EC from CTR, CUMS and VOR group; (B) Tilted frontal-side view of brain regions whose glucose metabolism altered; (C-G) Glucose metabolism was markedly decreased in CUMS group compared with CTR in bilateral mPFC (C), STR (D), AMY (E), HIP (F) and EC (G), while vortioxetine treatment restored glucose metabolism of right mPFC (C), AMY (E), HIP (F) and EC (G) in CUMS group. Results are showed as means ± S.D. (n = 7 per group, *p < 0.05, **p < 0.01, ***p < 0.001)

### 3.4 mRNA level of Cytokine in hippocampus, frontal cortex and striatum

Hippocampus, frontal cortex and striatum mRNA levels of YY1, IL-6, IL-1β and NF-kB-mRNA levels were measured in different groups by qPCR. Firstly, in frontal cortex area, we did detected significant increase of IL-1β (p=0.0033, p=0.0026, respectively) and YY1-mRNA (p=0.0321, p=0.0177, respectively) changes in CUMS than CTR or VOR group (Fig 4A-D). As for striatum, CUMS groups showed siginificantly increased of IL-1β (p=0.0079, p=0.0051, respectively, Fig 4E), YY1-mRNA expression compared to CTR or VOR group (p=0.0389, p=0.0002, respectively, Fig 4H), NF-kB-mRNA expression was elevated in CUMS than CTR, whereas do not descend after VOR treatment (p=0.0148, p=0.4170, respectively, Fig 4G). In hippocampus the IL-1β (p=0.0057, p=0.0017, respectively), NF-kB (p=0.0170, p=0.0362, respectively) and YY1-mRNA(p=0.0102, p=0.0017, respectively) was significant increase in CUMS in comparison with control or VOR group (Figure 4I, K, L), IL-6-mRNA expression level was also elevated in CUMS than control, while Vortioxetine treated showed no significant changes (p=0.0442, p=0.0697, respectively, Figure 4J).

**Figure 4.**
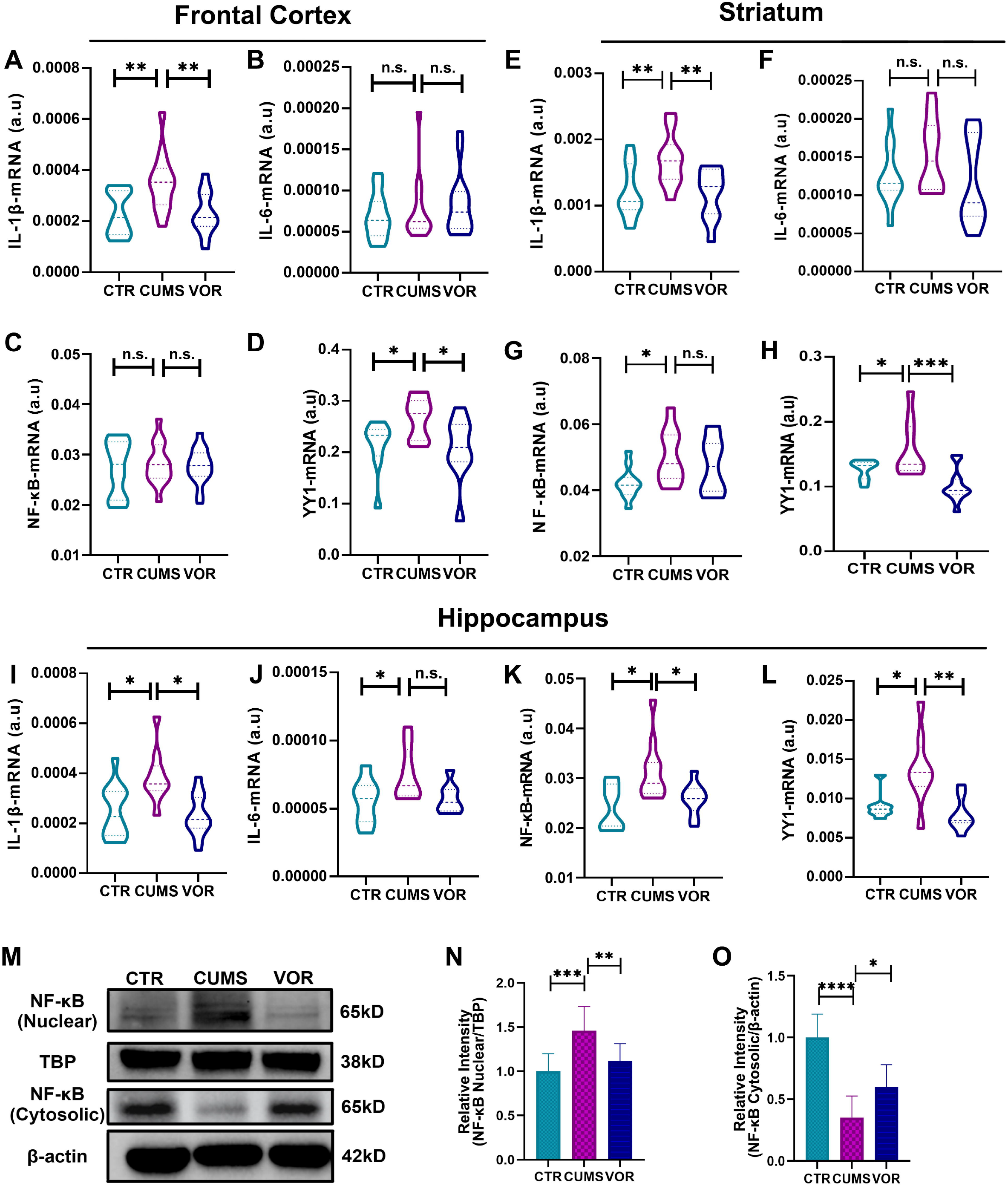
Changes of IL-1β, IL-6, NF-kB and YY1-mRNA expression in Frontal cortex (A-D), Striatum (E-H), hippocampus (I-L) among CTR (n = 8), CUMS (n = 8) and VOR group (n = 8); (M) Representative western blot image of NF-kB in nuclear level and cytosolic in the hippocampus among CTR, CUMS and VOR groups; (N) The ratio of nuclear NF-kB/nuclear TBP; (O) The ratio of cytosolic NF-kB/cytosolic β-actin) (n = 8 per group, *p < 0.05, **p < 0.01, ***p < 0.001, n.s.: non-significant difference)

### 3.5 Protein level of NF-κB in hippocampus

Since NF-κB may be an important mediator of YY1 regulation of inflammation, we investigated the protein expression at nuclear levels (Fig 4N) cytoplasmic (Fig 4O). The cytoplasmic protein level of NF-κB was significantly decreased in the CUMS group either than CTR or VOR group (p<0.0001, p=0.026, respectively), whereas the nuclear NF-κB level elevated in the CUMS group comparing to control and VOR treatment group (p=0.0006, p=0.0064, respectively).

## 4. Discussion

In the present study, the role of YY1 in depression associated inflammation and cognitive impairment was investigated in MDD patients and CUMS rodent model. The results showed increased plasma level of YY1 combined with cognitive impairment and elevated IL-1β and IL-6 level in MDD patients, which could be restored by vortioxetine treatment. Meanwhile, the result of rodents study is consistent with clinical findings that CUMS rats showed depressive and cognitive impairment which were reversed by vortioxetine administration. Among related depressive and connition brain area ie: frontal cortex, hippocamus, striatum, Increased hippocampus YY1, NF-κB, IL-6 and IL-1β in CUMS rats, which were suppressed by treated with vortioxetine. Protein level of NF-κB was increased in the nuclear level of CUMS than control and VOR group in hippocampus, while cytosolic level changes in the opposite way.^18^F-FDG microPET/CT scanning revealed decreased glucose uptake in hippocampus, amygdala, striatum and entorhinal cortex of CUMS group which reserved by vortioxetine.

Notably, both MDD patients and CUMS model rats showed recovery in mood and cognitive function after treatment with vortioxetine, which suggests that there may be common underlying mechanisms between depression and cognitive impairment. In fact, the structures of the medial temporal lobe, including the amygdala, hippocampus and entorhinal cortex, are involved in a variety of tasks, such as the production of emotional behaviors, the formation and storage of emotional memories, the consolidation of explicit memories of emotional arousal events(Bombardi, 2014, LeDoux, 2007, Eichenbaum and Lipton, 2008), and spatial information processing and memory, which is consistent with the patient’s clinical symptoms and our findings in animals. In our study, ^18^F-FDG scanning indicated decreased glucose uptake in hippocampus, amygdala and entorhinal cortex of CUMS group rats, and they increased after subsequent treatment. Many previous studies have shown significant changes in glucose metabolism in several brain regions in MDD patients, but the results have been inconsistent due to confounding factors such as age, medication and course of disease. Mayberg et al. reported reduced glucose metabolism in the frontal, hypothalamus, and medial frontal cortex(Hendler et al., 1997). Meanwhile, decreased brain metabolism in cingulate gyrus, superior frontal gyrus, rectum gyrus and orbital gyrus was also observed in MDD patients(Wei et al., 2016). The antidepressant effects of electric shock therapy were associated with increased metabolism in the left subgenicular anterior cingulate gyrus and hippocampus(McCormick et al., 2007). Recent studies have shown that brain hypometabolic pattern is closely related to the type and severity of cognitive deficits(Laforce et al., 2018). Previous study also demonstrated that chronic stress could lead to decreased glucose metabolism in brain regions of rats, such as amygdala and hippocampus (Wei et al., 2018, Van Laeken et al., 2018). Typhoid vaccination caused inflammation in healthy volunteers, which led to a sharp drop in MTL glucose metabolism and selectively impaired human spatial memory, which is similar to what we found in animals.

We also found changes in the levels of IL-1β and YY1 in peripheral blood of depressed patients before and after treatment, and positively correlation between YY1 and IL-1β, whereas, the same changes were found only in the hippocampus of the three available animal brain regions both in the mRNA changes and glucose metabolism level, therefore, the inflammatory mediated by the hippocampal YY1-NF-κB-IL-1β pathway is at least partially involved in the improvement of cognition and mood in depressed individuals by vortioxetine. A previous study has found the level of TNF-α, IL-17, IL-6, and IL-1β were markedly decreased in YY1-deficient mice with collagen-induced arthritis(Kwon et al., 2018). YY1 was upregulated in lipopolysaccharide (LPS) -stimulated BV2 microglia dependent on NF-κB activity. Furthermore, YY1 knockdown also inhibited LPS-induced the activation of NF-κB signaling BV-2□cells. Common binding sequences of NF-κB have been identified in promoter regions of several cytokine genes, including TNF-α and IL-1β(Muller et al., 1993), and inhibition of NF-κB inhibits the synthesis and release of IL-1β and TNF-α(Kuo et al., 2000). Though in vitro experiments on BV-2 cells showed that YY1 knockdown significantly inhibited LPS-induced IL-6 levels but not IL-1β(Zhang et al., 2018), and YY1 positively regulated IL-6 transcription by binding to the promoter region of the IL-6 gene as well(Lin et al., 2017), nonetheless, our results indicated that IL-1β appears to play a more prominent role in the treatment of depressed individuals with vortioxetine. Actually, brain IL-1β is necessary for depressive symptoms induced by chronic mild stress, and chronic IL-1β exposure also induces depressive behavior and reduces hippocampal neurogenesis(Goshen et al., 2008). Menin deficiency increases NF-κB-induced IL-1β levels in astrocytes, contributing to depression-like behavior(Leng et al., 2018). Il-1β knockdown in the hippocampus significantly alleviated IPS-induced memory deficits and anxiety- and depression-like behaviors in mice(Li et al., 2017). In clinical studies, the use of endotoxin in healthy volunteers resulted in increased levels of anxiety and depression in subjects, which is related to the transcription factor NF-κB(Cho et al., 2019).

This is a multi-dimensional study that comprehensively illustrates the role of YY1/NF-κB mediated inflammatory pathway in depression. But the study still has some limitations. Firstly, this study is an aggregated crossover study, but we did not verify the findings in rats with depression in human brain. Additional evidence may be needed to support the role of YY1/NF-κB -mediated inflammatory pathway in cognitive function and mood changes in depression, which may affect the overall interpretation of our study results. Secondly, the representative sample size of clinical samples is unknown, and our patients are mainly outpatients and inpatients, which may limit the universality of our study results. Thirdly, the relatively small sample size of this study may result in some potential relationships not being discovered. In the future, multi-center studies should be established to test the generality of these findings by recruiting more patients with different severity levels for long-term follow-up. Forthly, we have not further explored how inflammation causes depression-like behavior and cognitive impairment, and though studies have shown that depression patients have increased inflammation in both central and peripheral locations, the specific mechanism and link between them may need to be explored further in subsequent experiments. Finally, depression is a very heterogeneous disease. We did not classify patients with depression according to the characteristics of depression (including anhedonia and suicidal ideation), but only focused on the characteristics of cognitive impairment of these patients, and we included patients with moderate or severe depression. The lack of a more specific grouping may make the statistical results of our study less convincing to a certain extent. In addition, we only constructed the CUMS model, and did not verify such changes in other depression-like models such as chronic immobilization stress model and chronic social defeat stress.

Overall, our study suggests that YY1-NF-κB -IL-1β inflammatory pathway may play an essential part in vortioxetine’s effect on mood change and cognitive improvement in depression, and may be associated with changes in glucose metabolism in the mood regulation and cognitive related brain regions. These findings provide new evidence for the inflammatory mechanisms of depression and cognition repairment, and this multi-dimensional study may be beneficial to the development of diagnostic or therapeutic tools for depression in the future.

## Data Availability

All data produced in the present work are contained in the manuscript

## Acknowledgements

The figure of the article was drawn by Figdraw (www.figdraw.com). We sincerely thank the support of funds from the Medical Science and Technology Project of Zhejiang Province (2022RC024) and Natural Science Foundation of Zhejiang Province (LQ20H090010).

